# Understanding ethnic diversity in open dementia neuroimaging datasets

**DOI:** 10.1101/2023.04.27.23289208

**Authors:** Nicholas Heng, Timothy Rittman

## Abstract

**Introduction:** Ethnic differences in dementia are increasingly recognised in epidemiological measures and diagnostic biomarkers. Nonetheless, ethnic diversity remains limited in many study populations Here we provide insights into ethnic diversity in open access neuroimaging dementia datasets.

**Methods:** Datasets comprising dementia populations who underwent neuroimaging assessment with available data on ethnicity were included. Statistical analyses of sample and effect sizes were based on the Cochrane Handbook.

**Results:** 14 databases were included, with 12 studies of healthy and MCI groups, and 11 of dementia groups. Combining all studies, the largest ethnic group was Caucasian (21,512 participants) with the next most common being Afro-Caribbean (1,960), followed by Asian (780). The smallest effect size detectable within the Caucasian group was 0.03, compared to Afro-Caribbean (0.1) and Asian (0.16).

**Discussion:** Our findings quantify the lack of ethnic diversity in openly available neuroimaging dementia datasets. More representative data would facilitate the development and validation of neuroimaging biomarkers relevant across ethnicities.

## Introduction

The past few decades have seen growing interest in the field of biomarkers for neurodegenerative conditions. The neuroimaging community has led the way in open data ^1^, facilitating an explosion of research in neuroimaging biomarkers for dementia ^2^. This interest is in the context of an increasing global burden of neurodegenerative disorders, particularly in relation to the impact of Alzheimer’s disease and other dementias on an increasingly ageing population ^3^. Crucially, it has been estimated that the prevalence of dementia will increase from 57.4 million cases globally in 2019 to 152.8 million cases in 2050 ^4^, posing a considerable risk to global healthcare and society in the near future.

There has been emerging evidence of ethnic differences amongst dementia populations, not only in prevalence and incidence, but also in cerebrospinal fluid (CSF) and imaging biomarkers ^5-6^. Nonetheless, many studies remain homogenous in the ethnicity of participants ^7^. This may hinder the translation of results to real world applications. As such, we aimed to provide insights into the ethnic diversity of currently available open neuroimaging dementia databases worldwide.

## Materials and methods

We compiled and analysed demographic data reported by open access neuroimaging dementia databases. Databases were included if they consisted of (a) patients with a diagnosis of dementia or mild cognitive impairment, (b) had available neuroimaging data, and (c) demographic data including the breakdown of ethnicities. Datasets were identified through online research platforms including the Global Alzheimer’s Association Interactive Network (GAAIN) (https://www.gaain.org/), individual database repositories, and via peer-reviewed journal articles. We excluded datasets of solely genetic forms of dementia since these may be associated with specific ethnicities, or include large families that might bias the estimate of the distribution of ethnicities. A total of 46 databases were found but 32 were subsequently excluded as they either only included healthy controls, had no available data on demographics, or did not include neuroimaging data. Given the different definitions of ethnicities available, we took a pragmatic approach using the most widely used terms in the literature that permitted comparison between studies.

Statistical analyses on combined mean and standard deviation was performed as laid out by the Cochrane Handbook ^8^, and effect sizes calculations were using the pwr package in R (version 4.2.2) with ^9^. To compare samples of presumed equal sizes, we performed a power calculation for a two sample t test, estimating the effect size or sample size detectable with 90% power at a significance level (p value) of 0.05. Sample sizes were initially computed by setting a range of effect sizes, while minimum detectable effect sizes for single ethnic groups were then calculated using the aggregated dementia patient populations of different ethnicities from the open access dementia databases.

## Data availability

Data sharing is not applicable to this article as no new data were created or analysed in this study.

## Results

### Demographics of dementia databases

A total of 14 dementia neuroimaging datasets were included, separated into the three diagnostic groups, with 12 including healthy participants (Supplementary Table 1), 12 including patients with mild cognitive impairment (MCI) (Supplementary Table 2) and 11 including patients with dementia (Table 1) ^10-22^. In these tables, two entries for the Alzheimers’ Disease Neuroimaging Initiative (ADNI) dataset were made due to the separation of the ADNI-1 from ADNIGO and ADNI-2 cohorts. The majority of patients were from North America and Europe, with the two largest databases being from the National Alzheimer’s Coordinating Center (NACC) and UK Biobank respectively, in which there were a considerably higher percentage of Caucasians compared to other ethnicities.

**Table 1.**
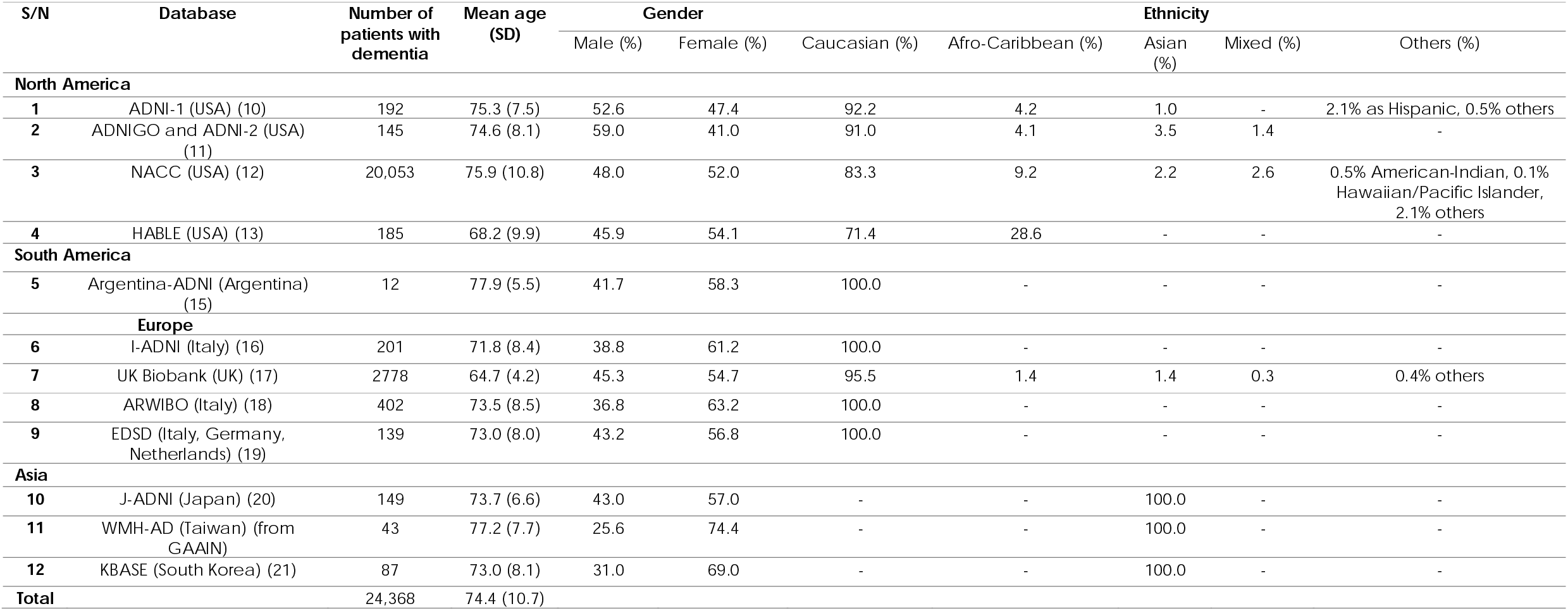
Table showing the breakdown of demographics data of patients with dementia in databases globally as separated by region.

### Effect size analyses

To understand how the breakdown of ethnicity in these datasets could affect research studies, we calculated the sample sizes required for a range of effect sizes. For example, based on a recent systematic review and meta-analysis on fluid biomarkers for Alzheimer’s disease ^6^, it was found that CSF p-tau_181_ and t-tau levels were significantly higher in the Caucasian population compared to African Americans with MCI, with a standard mean difference of -0.50 (95%CI -0.73 to -0.28) and -0.52 (95%CI -0.75 to -0.30) respectively – though bearing in mind these did not necessarily inform the effect size in other biomarkers or ethnicities. Therefore, using an estimated effect size of 0.50 and basing off a power calculation of 90% and significance level of 0.05, the number of patients required to detect a difference was N=86 each for two groups of patients of different ethnicities. We went on to calculate sample sizes for a range of effect sizes to obtain a better idea of the sample size to consider when planning future studies. In addition, we assessed whether the available data were sufficient to make comparisons between the Caucasian population and other ethnic groups (Table 2).

**Table 2.**
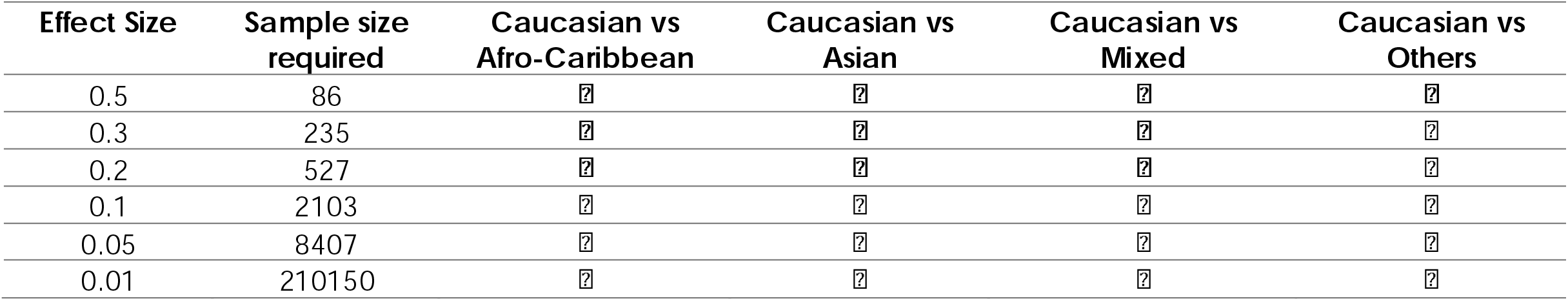
**Table showing the sample sizes required for specific effect sizes to be obtained based on power calculations of 90% and significance level of 0.05, with subsequent columns showing whether comparisons between ethnic groups can be performed based on currently available data.**

In an alternate approach, using available data for patients with dementia in those datasets combined with similar power calculation of 90% and significance of 0.05, we determined the smallest detectable effect size given currently available data (Table 3). The Caucasian population had the smallest minimum detectable effect size at 0.03 due to its size.

**Table 3.** Table showing the smallest effect sizes detected using population sizes based on available ethnicity data from dementia databases with similar parameters of 90% power and significance level of 0.05.

| Ethnicity group | Total number of patients with dementia | Smallest effect size detected |
| --- | --- | --- |
| Caucasian | 21512 | 0.03 |
| Afro-Caribbean | 1960 | 0.10 |
| Asian | 780 | 0.16 |
| Mixed | 532 | 0.20 |
| Others | 138 | 0.39 |

## Discussion

With the increasing number of studies focusing on ethnic differences in dementia, there is little doubt that more emphasis needs to be placed on the role that ethnic differences play in biomarker research. Our findings suggest that despite the vast amount of comprehensive and high-quality data available worldwide, most participants come from a Caucasian background, limiting comparison to other populations. Considerable numbers of patients are required for assessing small magnitude effect sizes – which becomes particularly important when trying to identify potentially subtle differences between ethnicities. The minimum detectable effect size can therefore act as a guide or threshold towards that end. In fact, the majority of the population sizes were made up of two large databases in the UK and US. We hope these findings can act as a starting point into deciding how to expand representation of different ethnic groups in future studies on dementia.

Understanding the limitations of currently available data can provide an opportunity to uncover and tackle the challenges associated with ensuring ethnic diversity in studies – stemming from a range of barriers to participation including literacy and language barriers, lack of understanding or misperceptions of the condition and services available, and cultural factors leading to mistrust of services or stigma surrounding the condition ^23^. There are some encouraging initiatives to address these challenges, such as those by the UK Dementia Research Institute, which are designed not only to facilitate recruitment of diverse patient cohorts by increasing awareness and broadening inclusion criteria, but also to improve access to healthcare through collaborations with local teams ^24^.

We were only able to obtain data for openly available datasets. We know from published data and from personal contacts that many studies use local cohorts, and some large national cohorts are not shared with the wider community. We advocate exploring the barriers to sharing those data, including the concerns of those who have collected and curate those datasets.

There are several limitations to our study – the first being that we were unable to comment on the representativeness (as opposed to heterogeneity) of the combined characteristics of the populations included in the database. Data on global ethnicity is not readily available and classifications differ between different countries, making it difficult to draw comparisons. Secondly, a considerable number of studies were excluded due to the lack of available demographic data, and those that were included were mainly based in the Western hemisphere, which may mean we have underestimate the non-Caucasian ethnicities actually available.

In our analysis we assume that datasets can easily be combined. In fact, harmonisation between datasets presents a significant methodological challenge given that protocols differ and site effects need to be modelled ^25-26^. This is particularly a challenge for combining neuroimaging data despite the increasing availability of tools for this purpose such as ComBat ^27^.

## Conclusion

With increasing awareness of the differences between ethnicities in dementia, it is imperative that we begin to prioritise and broaden research to better understand underlying mechanisms, to address the challenges associated with ethnic diversity in studies, and ultimately to pave the way for reliable translation into clinical practice.

## Data Availability

Data availability is not applicable to this article as no new data were created or analysed in this study.

https://www.gaain.org/

## Supplementary Materials

Supplementary tables 1 and 2 will be available online.

## References

1. Poldrack R., Gorgolewski K. Making big data open: data sharing in neuroimaging. Nat Neurosci 17, 1510–1517 (2014). https://doi.org/10.1038/nn.3818

2. Rittman T. Neurological update: neuroimaging in dementia. J Neurol. 2020 Nov;267(11):3429–3435. doi: 10.1007/s00415-020-10040-0. Epub 2020 Jul 7. PMID: 32638104; PMCID: PMC7578138.

3. GBD 2016 Neurology Collaborators. Global, regional, and national burden of neurological disorders, 1990–2016: a systematic analysis for the Global Burden of Disease Study 2016. Lancet Neurol. 2019; 18(5): 459–480.

4. GBD 2019 Dementia Forecasting Collaborators. Estimation of the global prevalence of dementia in 2019 and forecasted prevalence in 2050: an analysis for the Global Burden of Disease Study 2019. Lancet Public Health. 2022; 7(2): e105–e125.

5. Wilkins CH, Windon CC, Dilworth-Anderson P et al. Racial and Ethnic Differences in Amyloid PET Positivity in Individuals With Mild Cognitive Impairment or Dementia: A Secondary Analysis of the Imaging Dementia-Evidence for Amyloid Scanning (IDEAS) Cohort Study. JAMA Neurol. 2022; 79(11): 1139–1147.

6. Chaudhry A and Rizig M. Comparing fluid biomarkers of Alzheimer’s disease between African American or Black African and white groups: A systematic review and meta-analysis. J Neurol Sci. 2021; 421: 117270.

7. Vyas MV, Raval PK, Watt JA, Tang-Wai DF. Representation of ethnic groups in dementia trials: systematic review and meta-analysis. J Neurol Sci. 2018; 394: 107–111.

8. Higgins JPT, Green S: Cochrane Handbook for Systematic Reviews of Interventions. Wiley Online Library. 2008.

9. R Core Team. R: A language and environment for statistical computing. R Foundation for Statistical Computing, Vienna, Austria. 2013. Available from: http://www.R-project.org/.

10. Petersen RC, Aisen PS, Beckett LA, Donohue MC, Gamst AC, Harvey DJ, Jack CR, Jagust WJ, Shaw LM, Toga AW, Trojanowski JQ, Weiner MW. Alzheimer’s Disease Neuroimaging Initiative (ADNI). Neurology. 2010; 74(3): 201–209.

11. Aisen PS, Petersen RC, Donohue M, Weiner MW. ADNI 2 Clinical Core: Progress and Plans. Alzheimers Dement. 2015; 11(7): 734–739.

12. The NIA Alzheimer’s Disease Research Centers Program. National Alzheimer’s Coordinating Center. 2023. Available from: https://naccdata.org/requesting-data/data-summary/uds.

13. O’Bryant SE, Johnson LA, Barber RC, Braskie MN, Christian B, Hall JR, Hazra N, King K, Kothapalli D, Large S, Mason D, Matsiyevskiy E, McColl R, Nandy R, Palmer R, Petersen M, Philips N, Rissman RA, Shi Y, Toga AW, Vintimilla R, Vig R, Zhang F, Yaffe K; HABLE Study Team. The Health & Aging Brain among Latino Elders (HABLE) study methods and participant characteristics. Alzheimers Dement (Amst). 2021;13(1):e12202.

14. LaMontague PJ, Benzinger TLS, Morris JC, Keefe S, Hornbeck R, Xiong C, Grant E, Hassenstab J, Moulder K, Vlassenko AG, Raichile ME, Cruchaga C, Marcus D. OASIS-3: Longitudinal Neuroimaging, Clinical, and Cognitive Dataset for Normal Aging and Alzheimer Disease. 2019. medRxiv. doi: 10.1101/2019.12.13.19014902.

15. Méndez PC, Calandri I, Nahas F, Russo MJ, Demey I, Martín ME, Clarens MF, Harris P, Tapajoz F, Campos J, Surace EI, Martinetto H, Ventrice F, Cohen G, Vázquez S, Romero C, Guinjoan S, Allegri RF, Sevlever G. Argentina-Alzheimer’s disease neuroimaging initiative (Arg-ADNI): neuropsychological evolution profile after one-year follow up. Arq Neuropsiquiatr. 2018;76(4):231–240.

16. Cavedo E, Redolfi A, Angeloni F, Babiloni C, Lizio R, Chiapparini L, Bruzzone MG, Aquino D, Sabatini U, Alesiani M, Cherubini A, Salvatore E, Soricelli A, Vernieri F, Scrascia F, Sinforiani E, Chiarati P, Bastianello S, Montella P, Corbo D, Tedeschi G, Marino S, Baglieri A, De Salvo S, Carducci F, Quattrocchi CC, Cobelli M, Frisoni GB. The Italian Alzheimer’s Disease Neuroimaging Initiative (I-ADNI): validation of structural MR imaging. J Alzheimers Dis. 2014;40(4):941–52.

17. Swaddiwudhipong N, Whiteside DJ, Hezemans FH, Street D, Rowe JB, Rittman T. Pre-diagnostic cognitive and functional impairment in multiple sporadic neurodegenerative diseases. Alzheimers Dement. 2022.

18. NeuGRID2 consortium. NeuGRID. 2012. Available from: https://www.neugrid2.eu/index.php/introduction/

19. Brueggen K, Grothe MJ, Dyrba M, Fellgiebel A, Fischer F, Filippi M, Agosta F, Nestor P, Meisenzahl E, Blautzik J, Frölich L, Hausner L, Bokde ALW, Frisoni G, Pievani M, Klöppel S, Prvulovic D, Barkhof F, Pouwels PJW, Schröder J, Hampel H, Hauenstein K, Teipel S. The European DTI Study on Dementia - A multicenter DTI and MRI study on Alzheimer’s disease and Mild Cognitive Impairment. Neuroimage. 2017;144(Pt B):305–308.

20. NBDC Human Database. NBDC Research ID: hum0043.v1. 2016. Available from: https://humandbs.biosciencedbc.jp/en/hum0043-v1

21. Byun MS, Yi D, Lee JH, Choe YM, Sohn BK, Lee JY, Choi HJ, Baek H, Kim YK, Lee YS, Sohn CH, Mook-Jung I, Choi M, Lee YJ, Lee DW, Ryu SH, Kim SG, Kim JW, Woo JI, Lee DY; KBASE Research Group. Korean Brain Aging Study for the Early Diagnosis and Prediction of Alzheimer’s Disease: Methodology and Baseline Sample Characteristics. Psychiatry Investig. 2017;14(6):851–863.

22. Albani D, Marizzoni M, Ferrari C, Fusco F, Boeri L, Raimondi I, Jovicich J, Babiloni C, Soricelli A, Lizio R, Galluzzi S, Cavaliere L, Didic M, Schönknecht P, Molinuevo JL, Nobili F, Parnetti L, Payoux P, Bocchio L, Salvatore M, Rossini PM, Tsolaki M, Visser PJ, Richardson JC, Wiltfang J, Bordet R, Blin O, Forloni G, Frisoni GB; PharmaCog Consortium. Plasma Aβ42 as a Biomarker of Prodromal Alzheimer’s Disease Progression in Patients with Amnestic Mild Cognitive Impairment: Evidence from the PharmaCog/E-ADNI Study. J Alzheimers Dis. 2019;69(1):37–48. doi: 10.3233/JAD-180321. PMID: 30149449.

23. Kenning C, Daker-White G, Blakemore A, Panagioti M, Waheed W. Barriers and facilitators in accessing dementia care by ethnic minority groups: a meta-synthesis of qualitative studies. BMC Psychiatry. 2017; 316.

24. UK Dementia Research Institute. Diversity and dementia: how is research reducing health disparities? 2022. https://ukdri.ac.uk/uploads/UK-DRI_Dementia_Health_Inequalities_Report_2022.pdf

25. Lipnicki DM, Lam BCP, Mewton L, Crawford JD, Sachdev PS. Harmonizing Ethno-Regionally Diverse Datasets to Advance the Global Epidemiology of Dementia. Clin Geriatr Med. 2023 Feb;39(1):177–190. doi: 10.1016/j.cger.2022.07.009. Epub 2022 Oct 18. PMID: 36404030; PMCID: PMC9767705.

26. Shishegar R, Cox T, Rolls D, Bourgeat P, Doré V, Lamb F, Robertson J, Laws SM, Porter T, Fripp J, Tosun D, Maruff P, Savage G, Rowe CC, Masters CL, Weiner MW, Villemagne VL, Burnham SC. Using imputation to provide harmonized longitudinal measures of cognition across AIBL and ADNI. Sci Rep. 2021 Dec 10;11(1):23788. doi: 10.1038/s41598-021-02827-6. PMID: 34893624; PMCID: PMC8664816.

27. Pomponio R, Erus G, Habes M, Doshi J, Srinivasan D, Mamourian E, Bashyam V, Nasrallah IM, Satterthwaite TD, Fan Y, Launer LJ, Masters CL, Maruff P, Zhuo C, Völzke H, Johnson SC, Fripp J, Koutsouleris N, Wolf DH, Gur R, Gur R, Morris J, Albert MS, Grabe HJ, Resnick SM, Bryan RN, Wolk DA, Shinohara RT, Shou H, Davatzikos C. Harmonization of large MRI datasets for the analysis of brain imaging patterns throughout the lifespan. Neuroimage. 2020 Mar;208:116450. doi: 10.1016/j.neuroimage.2019.116450. Epub 2019 Dec 9. PMID: 31821869; PMCID: PMC6980790.

