## Supplementary material for "Understanding ethnic diversity in open dementia neuroimaging datasets": Supllementary Tables 1 and 2

**Supplementary Tables**

**Supplementary Table 1. Table showing the breakdown of demographics data of healthy controls or total numbers in databases globally as separated by region.**

| S/N | Database | Number of healthy controls | Mean age (SD) | Gender | | Ethnicity | | | | |
|---|---|---|---|---|---|---|---|---|---|---|
| | | | | Male (%) | Female (%) | Caucasian (%) | Afro-Caribbean (%) | Asian (%) | Mixed (%) | Others (%) |
| **North America** | | | | | | | | | | |
| **1** | ADNI-1 (USA) (10) | 229 | 75.8 (5.0) | 52.0 | 48.0 | 90.8 | 7.0 | 1.3 | - | 0.9% Hispanic |
| **2** | ADNIGO and ADNI-2 (USA) (11) | 287 | 73.0 (6.1) | 46.0 | 54.0 | 90.2 | 5.9 | 1.7 | 1.7 | 0.3% American-Indian/Alaskan |
| **3** | NACC (USA) (12) | 16,825 | 72.8 (11.4) | 35.0 | 65.0 | 76.8 | 14.8 | 3.1 | 3.4 | 0.6% American-Indian, 0.1% Hawaiian/Pacific Islander, 1.2% others |
| **4** | HABLE (USA) (13) | 2032 | 65.0 (8.4) | 34.9 | 65.1 | 84.0 | 15.9 | - | - | N=1 mixed |
| **5** | OASIS-3 (USA) (14) | 1098 | 68.8 (range 42.5-95.6) | 44.4 | 55.6 | 84.3 | 15.3 | 0.4 | - | - |
| **South America** | | | | | | | | | | |
| **6** | Argentina-ADNI (Argentina) (15) | 14 | 70.1 (8.2) | 28.6 | 71.4 | 100.0 | - | - | - | - |
| **Europe** | | | | | | | | | | |
| **7** | I-ADNI (Italy) (16) | 7 | 70.0 (10.5) | 57.1 | 42.9 | 100.0 | - | - | - | - |
| **8** | UK Biobank (UK) (17) | 493,735 | 56.4 (8.1) | 44.3 | 52.9 | 93.9 | 1.6 | 2.3 | 0.5 | 0.9% others |
| **9** | ARWIBO (Italy) (18) | 1482 | 51.6 (15.4) | 39.1 | 60.9 | 100.0 | - | - | - | - |
| **10** | EDSD (Italy, Germany, Netherlands) (19) | 194 | 69.0 (6.0) | 49.0 | 51.0 | 100.0 | - | - | - | - |
| **Asia** | | | | | | | | | | |
| **11** | J-ADNI (Japan) (20) | 154 | 68.3 (5.8) | 48.1 | 51.9 | - | - | 100.0 | - | - |
| **12** | WMH-AD (Taiwan) (From GAAIN) | 19 | 70.4 (8.2) | 68.4 | 31.6 | - | - | 100.0 | - | - |
| **13** | KBASE (South Korea) (21) | 365 (Total) | 62.8 (15.2) | 48.5 | 51.5 | - | - | 100.0 | - | - |
| **Total** | (Excluding OASIS-3 due to range) | 515,343 | 57.0 (8.8) | | | | | | | |

**Supplementary Table 2. Table showing the breakdown of demographics data of patients with mild cognitive impairment (MCI) in databases globally as separated by region.**

| S/N | Database | Number of patients with MCI | Mean age (SD) | Gender | | Ethnicity | | | | |
|---|---|---|---|---|---|---|---|---|---|---|
| | | | | Male | Female | Caucasian | Afro-Caribbean | Asian | Mixed | Others |
| **North America** | | | | | | | | | | |
| 1 | ADNI-1 (USA) (10) | 398 | 74.7 (7.4) | 64.6% | 35.4% | 90.5% | 3.5% | 2.3% | - | 3.5% classed as Hispanic, 0.3% as others |
| 2 | ADNIGO and ADNI-2 (USA) (11) | 461 | 71.6 (7.4) | 55.3% | 44.7% | 93.3% | 2.8% | 1.1% | 1.5% | 0.2% American-Indian/Alaskan, 0.4% Hawaiian/Pacific Islander, 0.7% others |
| 3 | NACC (USA) (12) | 8404 | 75.7 (10.2) | 47.0% | 53.0% | 75.6% | 15.4% | 3.0% | 3.5% | 0.7% American-Indian, 0.1% Hawaiian/Pacific Islander, 1.6% others |
| 4 | HABLE (USA) (13) | 490 | 65.9 (9.0) | 44.9% | 55.1% | 70.6% | 29.4% | - | - | - |
| **South America** | | | | | | | | | | |
| 5 | Argentina-ADNI (Argentina) (15) | 23 | 74.0 (7.4) | 47.9% | 52.1% | 100.0% | - | - | - | - |
| **Europe** | | | | | | | | | | |
| 6 | PharmaCog (Europe) (22) | 151 | 69.3 (7.4) | 43.7% | 56.3% | 100.0% | - | - | - | - |
| 7 | I-ADNI (Italy) (16) | 54 | 69.5 (7.2) | 50.0% | 50.0% | 100.0% | - | - | - | - |
| 8 | ARWIBO (Italy) (18) | 308 | 71.2 (8.0) | 38.3% | 61.7% | 100.0% | - | - | - | - |
| 9 | EDSD (Italy, Germany, Netherlands) (19) | 160 | 73.0 (7.0) | 56.9% | 43.1% | 100.0% | - | - | - | - |
| 10 | DCL (Spain) (From GAAIN and investigators) | 308 | 75.1 (8.7) | 51.3% | 48.7% | 100.0% | - | - | - | - |
| **Asia** | | | | | | | | | | |
| 11 | J-ADNI (Japan) (20) | 234 | 73.0 (5.9) | 49.6% | 50.4% | - | - | 100.0% | - | - |
| 12 | WMH-AD (Taiwan) (From GAAIN) | 27 | 73.4 (7.7) | 48.1% | 51.9% | - | - | 100.0% | - | - |
| 13 | KBASE (South Korea) (21) | 139 | 73.7 (7.0) | 33.8% | 66.2% | - | - | 100.0% | - | - |
| **Total** | | 11,157 | 74.7 (9.9) | | | | | | | |
